# Cytoplasmic staining of T cell receptor components enables efficient assessment of lineage and clonality in surface CD3-negative T cell neoplasms

**DOI:** 10.64898/2026.06.02.26354783

**Authors:** Aaron J. Wilk, Gary Gitana, Jean Oak

## Abstract

Flow cytometry can establish T cell clonality by detecting a restricted expression pattern of the T cell receptor (TCR) β constant region (TRBC), expressed in association with CD3. However, T cell neoplasms frequently lose surface expression of the CD3/TCR complex, posing a challenge to demonstrating T cell lineage and clonality. To address this challenge, here we present a 12-color flow cytometry panel, called cytoTCR, to characterize cytoplasmic expression of CD3/TCR complex components. We apply cytoTCR to 38 patient specimens with immunophenotypically abnormal T cell populations, demonstrating this approach can efficiently establish T cell lineage and clonality in challenging T cell neoplasms that have lost surface CD3 expression. While we show that natural killer (NK)-lineage neoplasms can express cytoplasmic CD3 at similar levels to T cells, we show that absent expression of cytoplasmic TCR components by mature lymphocytes can help confirm NK cell lineage. We demonstrate that cytoTCR can detect cytoplasmic TRBC-restriction in challenging cases of null-phenotype anaplastic large cell lymphoma, which lack surface expression of pan-T cell antigens. In cases of T-lymphoblastic leukemia, cytoTCR shows that cytoplasmic TRBC expression matches the expected developmental stage of the leukemia. Finally, we use cytoTCR to characterize atypical cCD3^-^CD7^-^ T cells in a patient with a history of T-lymphoblastic leukemia as well as recent CAR-T therapy, showing that this atypical population is polytypic and represents CAR-T product rather than residual disease. Our study presents a broadly applicable flow cytometric approach to simultaneously assess T cell lineage and clonality in suspected T lineage populations with absent surface CD3 expression.

## INTRODUCTION

T cell lineage is defined by somatic rearrangement of T cell receptor (TCR)–encoding genes, followed by expression of the CD3/TCR complex on the cell surface. In neoplasms of T cell origin, diagnosis typically involves three major objectives: (i) confirmation of T cell lineage, (ii) detailed phenotyping of T cell-associated antigens, and (iii) demonstration of clonality (Alaggio et al., 2022; WHO Classification of Tumours Editorial Board, 2024). When suitable specimens are available, flow cytometry is a powerful tool that can address each of these aims simultaneously (Oak et al., 2025).

For instance, flow cytometric analysis can identify candidate abnormal T cell populations based on antigenic aberrancy—either by detecting inappropriate loss of lineage-defining antigens such as CD3, CD2, CD5, or CD7, or by identifying inappropriate gain of markers, including bright expression of T cell antigens or acquisition of non-T cell antigens such as CD10 or CD30 (Horna et al., 2024a; Jevremovic & Olteanu, 2019; Kesler et al., 2007; Oak et al., 2025). More recently, the introduction of monoclonal antibodies specific to individual TCR β-chain constant regions (TRBC1 and TRBC2) has expanded the capability of flow cytometry to assess T cell clonality directly (Berg et al., 2021; Devitt et al., 2024; Horna et al., 2021, 2024a).

Only two β-chain constant regions exist, and incorporation of a β-chain constant region is required for productive rearrangement and expression of a TCRαβ receptor. Selection between TRBC1 and TRBC2 is a random, mutually exclusive event during T cell development (Horna et al., 2024a; Viney et al., 1992). Consequently, every mature αβ T cell should express one of TRBC1 or TRBC2, but never both. Surface staining for TRBC1 (with reciprocal inference of TRBC2 expression) therefore enables rapid detection of T cell clonality, as a clonal αβ T cell population will display restriction to a single TRBC isoform. Because surface expression of any CD3 subunit (except CD3ζ) requires assembly of the intact CD3/TCR complex, a CD3⁺ TCRγδ⁻ TRBC1⁻ population should by definition be TRBC2⁺ (Horna et al., 2024b). Thus, surface TRBC1 staining has emerged as a powerful and efficient method for establishing T cell clonality and is now increasingly incorporated into routine diagnostic flow cytometry panels.

A major limitation of TRBC-based clonality assessment, however, is that many T cell neoplasms—particularly those that are immature, dedifferentiated, or clinically aggressive—lose surface expression of the CD3/TCR complex (Alaggio et al., 2022; Hu et al., 2021; Kapur et al., 2014; Khodadoust & Silva, 2023; Oak et al., 2025). In such cases, conventional surface staining cannot reliably determine either lineage or clonality (**Figure 1A**). For instance, a cytotoxic T cell that has lost surface CD3 expression may appear immunophenotypically similar to an NK cell. Alternatively, null-phenotype anaplastic large cell lymphoma, which have absent surface expression of multiple T cell antigens, may appear immunophenotypically similar to monocytes by flow cytometry, though they are T cell lineage (**Figure 1A**). Historically, flow cytometry laboratories have relied on cytoplasmic CD3 (cCD3) staining to demonstrate T cell lineage in CD3-negative proliferations. Although cCD3 positivity often supports a T cell origin, it is not entirely specific because natural killer (NK) cells may show cytoplasmic expression of any CD3 subunit (including cCD3ε, cCD3γ, and cCD3δ) in the absence of TCR gene rearrangements and a functional TCR complex (Hao et al., 2021; Lanier et al., 1992; Wilk et al., 2021; Wu et al., 2021). Moreover, cCD3 staining does not directly assess clonality. While populations that are surface CD3-negative but cytoplasmic CD3-positive (sCD3⁻/cCD3⁺) are often presumed abnormal, distinguishing between neoplastic and non-neoplastic immature T cell proliferations—such as differentiating T-lymphoblastic leukemia (T-ALL) from indolent T-lymphoblastic proliferations—remains challenging.

**Figure 1.**
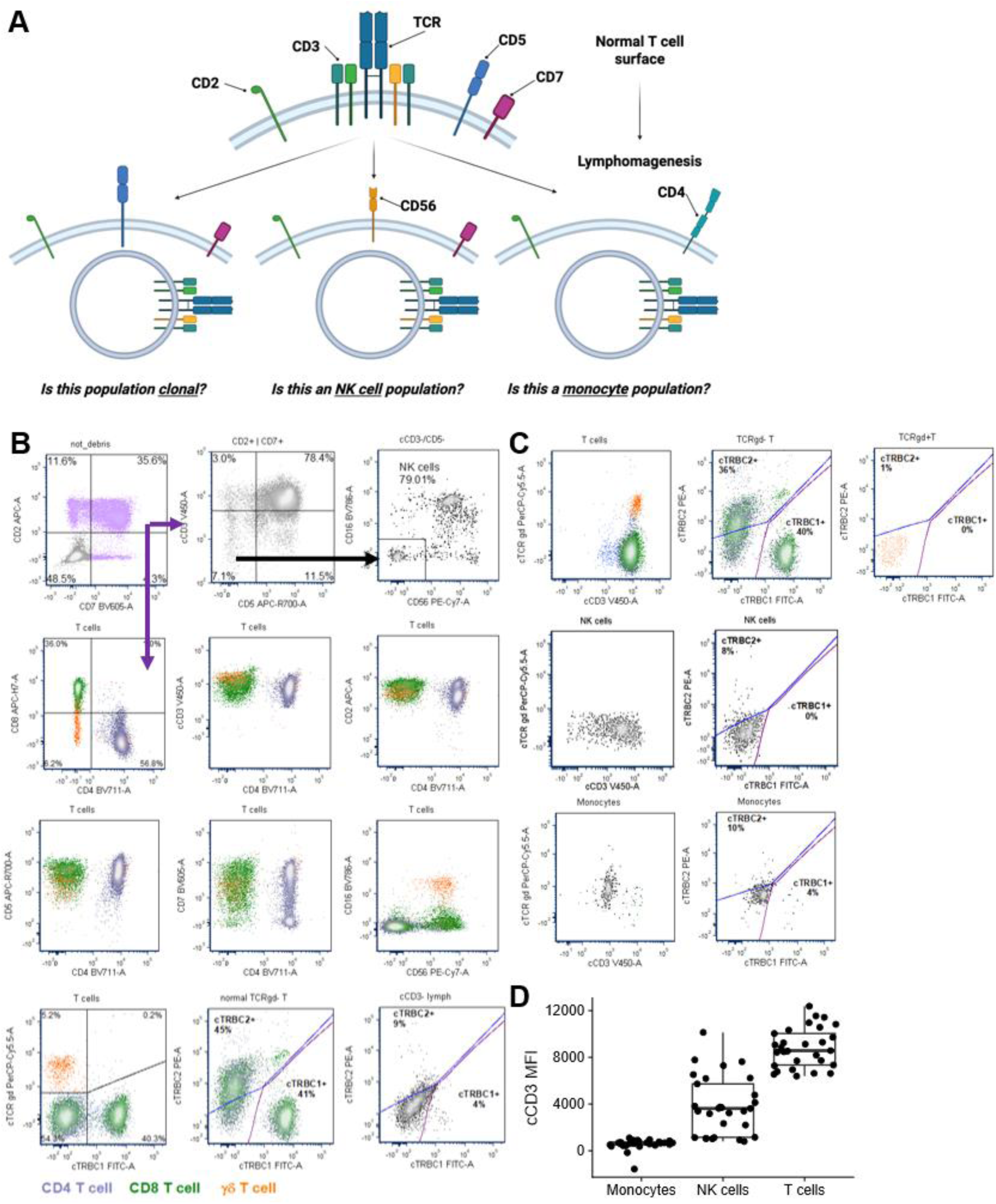
Gating scheme for cytoTCR panel. **A)** Illustration of utility of cytoplasmic staining of CD3/TCR complex components for surface CD3-negative candidate T cell populations. T lineage cells that have internalized or otherwise lost surface expression of the CD3/TCR complex cannot be directly assessed for clonality by surface staining alone, and may mimic cell types of other lineages, such as NK cells, or in rarer instances, monocytes. **B)** Gating scheme for cytoTCR. Candidate T cells are defined as any event positive for either CD2 or CD7. Next, NK cells are excluded by identifying cCD3^-^CD5^-^ events that express either CD16 and/or CD56. cTRBC1^+^ or cTRBC2^+^ T cells are identified using cCD3^-^ lymphocytes as a negative control. A normal peripheral blood specimen is shown demonstrating no abnormal antigen loss or gain and a representative distribution of cTRBC1, cTRBC2, and cTCRγδ. **C)** Distribution of cytoplasmic CD3, cTRBC1, cTRBC2, and cTCRγδ expression by T cells (top row), NK cells (middle row), and monocytes (bottom row). **D)** Boxplots depicting the mean fluorescent intensity (MFI) of cCD3 expression by monocytes, T cells, and NK cells in patients without a T or NK cell neoplasm.

To address these diagnostic challenges, we developed a multicolor intracellular flow cytometry panel, termed cytoTCR, which integrates cytoplasmic staining for all components of the CD3/TCR complex—CD3, TRBC1, TRBC2, and TCRγδ—with comprehensive surface immunophenotyping. This approach enables simultaneous evaluation of lineage, maturation stage, and clonality, in populations lacking surface CD3/TCR expression.

## MATERIALS & METHODS

### Patients and normal healthy donors

This study was approved by the Stanford University Institutional Review Board. Peripheral blood, bone marrow, tissue, and fluid specimens that were evaluated for standard leukemia/lymphoma immunophenotyping, including our surface T cell antigen panel (**Supplementary Table 1**), between January 7, 2024 and March 10, 2026, were included in our study. Peripheral blood specimens from healthy donors, which served as daily quality control for leukemia/lymphoma panels, were also included in our study over the same period as the control group. Patient demographic, pathologic, and molecular features were collected by chart review. Patients with documented histories of T cell neoplasms were also included in our study.

### Design and validation of cytoTCR

To design a panel for characterization of cytoplasmic CD3/TCR complex components, we modified our T cell surface antigen panel (**Supplementary Table 1**), by converting staining of all CD3/TCR complex components from surface staining to cytoplasmic staining, and adding cTRBC2 to this panel (**Table 1**). This panel therefore enables correlation of surface immunophenotypes between cytoTCR and our T cell surface antigen panel, while fully characterizing expression of the CD3/TCR complex via cytoplasmic staining for cCD3, cTRBC1, cTRBC2, and cTCRγδ.

**Table 1:**
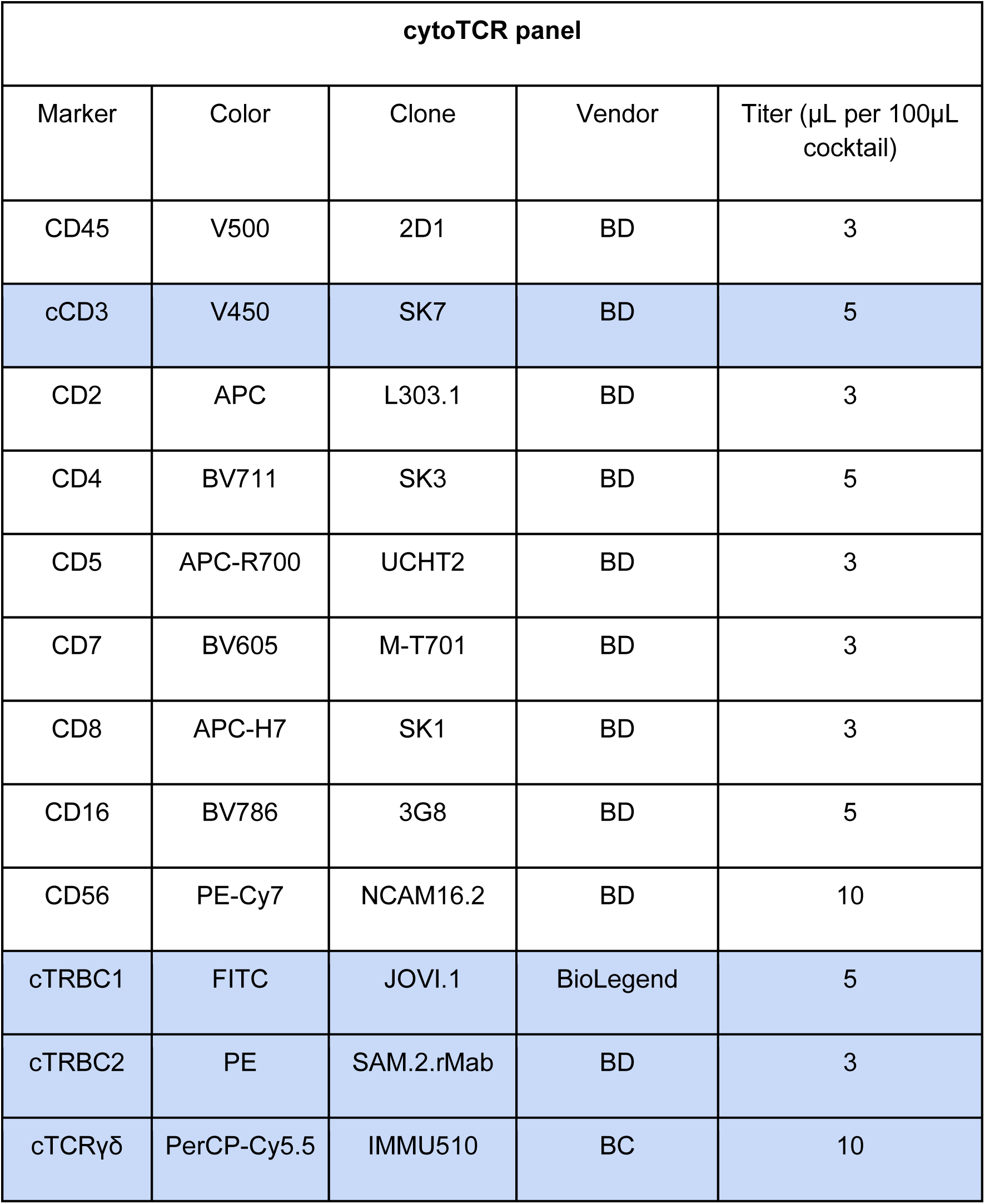
Antibody specificities, fluorophores, clones, vendors, and titers used for cytoTCR. Markers stained cytoplasmically are highlighted in blue.

### Flow cytometry immunophenotyping

Peripheral blood, bone marrow, and fluid specimens were first prepared using bulk lysis by ammonium chloride lyse solution. Next, 10 µL of Brilliant Stain buffer (BD Biosciences, San Jose, CA, USA) and the volumes of surface antibodies listed in **Table 1** were added to 100 µL of bulk lysed sample, followed by incubation at room temperature for 30 minutes. Next, samples were fixed and permeabilized using the Invitrogen FIX & PERM™ Cell Permeabilization Kit (Invitrogen, Carlsbad, CA, USA) according to the manufacturer’s specifications. Next, the volumes of cytoplasmic antibodies in **Table 1** were added and incubated at room temperature for 20 minutes, followed by one wash with DPBS before final resuspension in DPBS/0.1% sodium azide.

Specimens were analyzed by a Lyric 12-color flow cytometry instrument (BD Biosciences, San Jose, CA). Six instruments were in use over the time frame of this study. Data analyses were performed by using FCS Express Software (De Novo Software, Pasadena, CA). At least 200,000 cells were collected per tube. The following Boolean gating formulas were used to define candidate cell populations (**Figure 1B**):

T cells (T cell surface antigen tube): sCD3^+^ OR [(CD2^+^ OR CD7^+^) AND (NOT NK cells)] T cells (cytoTCR): (cCD3^+^ OR CD2^+^ OR CD7^+^) AND (NOT NK cells)

NK cells (T cell surface antigen tube): sCD3^-^ AND CD4^-^ AND CD5^-^ AND (CD2^+^ OR CD7^+^) AND (CD16^+^ OR CD56^+^)

NK cells (cytoTCR): CD4^-^ AND CD5^-^ AND (CD2^+^ OR CD7^+^) AND (CD16^+^ OR CD56^+^)

### Statistical analyses and visualization

Peripheral blood reference range verification for cTRBC1, cTRBC2, and cTCRγδ was performed using *n* = 21 peripheral blood specimens without evidence of clonal T cell population, using the non-parametric reference interval calculation implemented by the R package *referenceInterval* (Horowitz, 2008). Custom ggplot wrappers were used for all data visualizations other than flow cytometry data analysis. BioRender and LucidChart were utilized for diagrammatic illustrations.

## RESULTS

### A flow cytometry panel for characterization of suspected T cell populations lacking surface CD3

cytoTCR is designed to assess the cytoplasmic expression of all major components of the CD3/TCR complex, expression of which defines T cell lineage. This includes cytoplasmic staining of CD3, TRBC1, TRBC2, and TCRγδ (**Table 1**). Candidate T cells are first identified by gating all non-debris events positive for CD2 and/or CD7, followed by exclusion of NK cells (see **Methods**; **Figure 1B**). The resulting T cell population can then be evaluated for aberrant antigen gain/loss as well as for TCR monotypia. We applied cytoTCR to *n* = 21 peripheral blood specimens without evidence of monotypic T cell populations and demonstrated that cytoTCR yields comparable results of T cell subsets and TRBC1 distribution to surface staining (**Supplementary Figure 1**). From these data, we verified the non-parametric reference ranges for cTRBC1, cTRBC2, and cTCRγδ expression, which were compatible with previously reported TRBC and TCRγδ frequencies (**Supplementary Figure 1**) (Devitt et al., 2024; Oak et al., 2025).

cytoTCR recapitulates the biologically expected patterns of cytoplasmic CD3 and TCR component expression by various leukocyte subsets. For instance, cTCRγδ^+^ cells are negative for cTRBC1/2 (**Figure 1C**). Notably, we find that NK cells express cytoplasmic CD3 at levels comparable to slightly dim relative to background T cells but do not show cytoplasmic expression of any TCR components (**Figure 1C-D**; discussed further in **Figure 5**). Importantly, CD4^dim^ monocytes do not show nonspecific cytoplasmic expression of CD3 or any TCR component (**Figure 1C-D**).

### cytoTCR demonstrates T cell lineage and clonality in surface CD3⁻ T cell neoplasms

Next, we applied cytoTCR to specimens in which a surface CD3-negative suspected T cell population was identified. For instance, in a patient undergoing workup for persistent eosinophilia, a surface CD3-negative CD4^+^CD5^+^ lymphocyte population was detected by flow cytometry in peripheral blood (**Figure 2A**). cytoTCR was performed on this specimen to assess if the population represented T cell lineage, and if the population demonstrated TRBC-restriction. Indeed, cytoTCR demonstrated the population expressed cCD3 at a level similar to background T cells, and was monotypic for expression of cTRBC2 (**Figure 2B**). By simultaneously demonstrating the lineage and clonality of this population, cytoTCR enabled reporting of a TRBC-monotypic T cell population that, given the clinical history, was compatible with a diagnosis of lymphocyte-variant hypereosinophilic syndrome.

**Figure 2.**
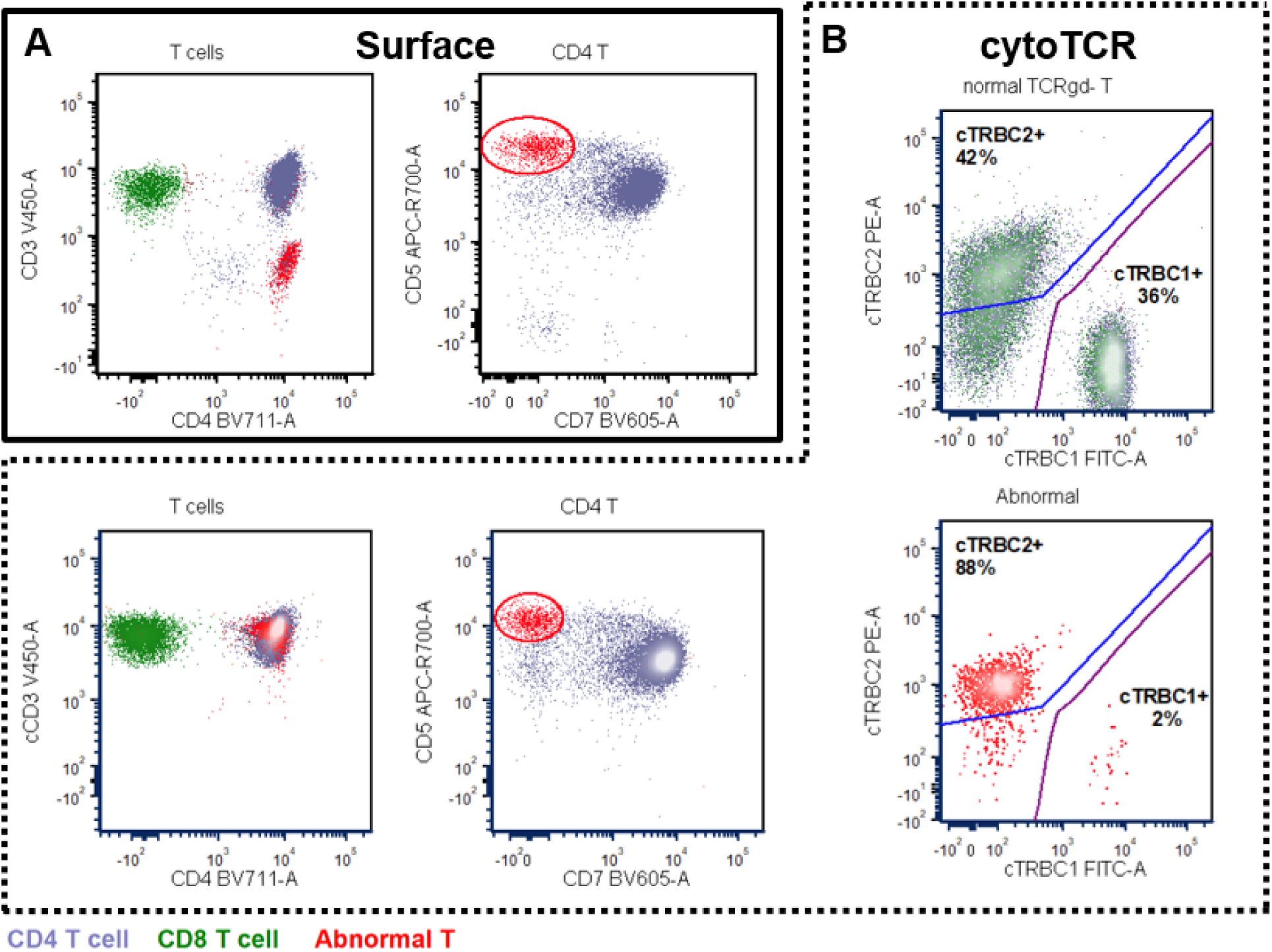
cytoTCR demonstrates clonality in surface CD3^-^ T cell populations. **A)** Standard T cell antigen surface staining identifies an abnormal surface CD3^-^ population expressing CD4 and CD5 in a patient undergoing workup for eosinophilia. **B)** cytoTCR demonstrates the abnormal population expresses cCD3 and is TRBC2-restricted, compatible with a diagnosis of lymphocyte-variant hypereosinophilic syndrome.

In total, we applied cytoTCR to 17 specimens with surface CD3^+^ TRBC-monotypic T cell populations, 16 suspected T-lineage populations lacking surface CD3, and 5 NK cell neoplasms (**Figure 3**). In all but one specimen with surface CD3^+^ monotypic T cell populations, the TRBC-restriction detected by cytoTCR matched the TRBC-restriction predicted by surface staining (**Figure 3A**). That is, sCD3^+^sTRBC1^-^sTCRγδ^-^ were found to be cTRBC2^+^. In cases where the monotypic T cell population demonstrated dim expression of TRBC1, the cytoplasmic TRBC restriction could be predicted by the intensity of surface CD3 expression as previously shown in other studies (**Figure 3A**) (Shi et al., 2025). In cases with dim surface TRBC1 expression, all populations were found to be cTRBC2-restricted with the exception of those with dim sCD3 expression, which were cTRBC1-restricted, matching prior reports (**Figure 3A**) (Shi et al., 2025).

**Figure 3.**
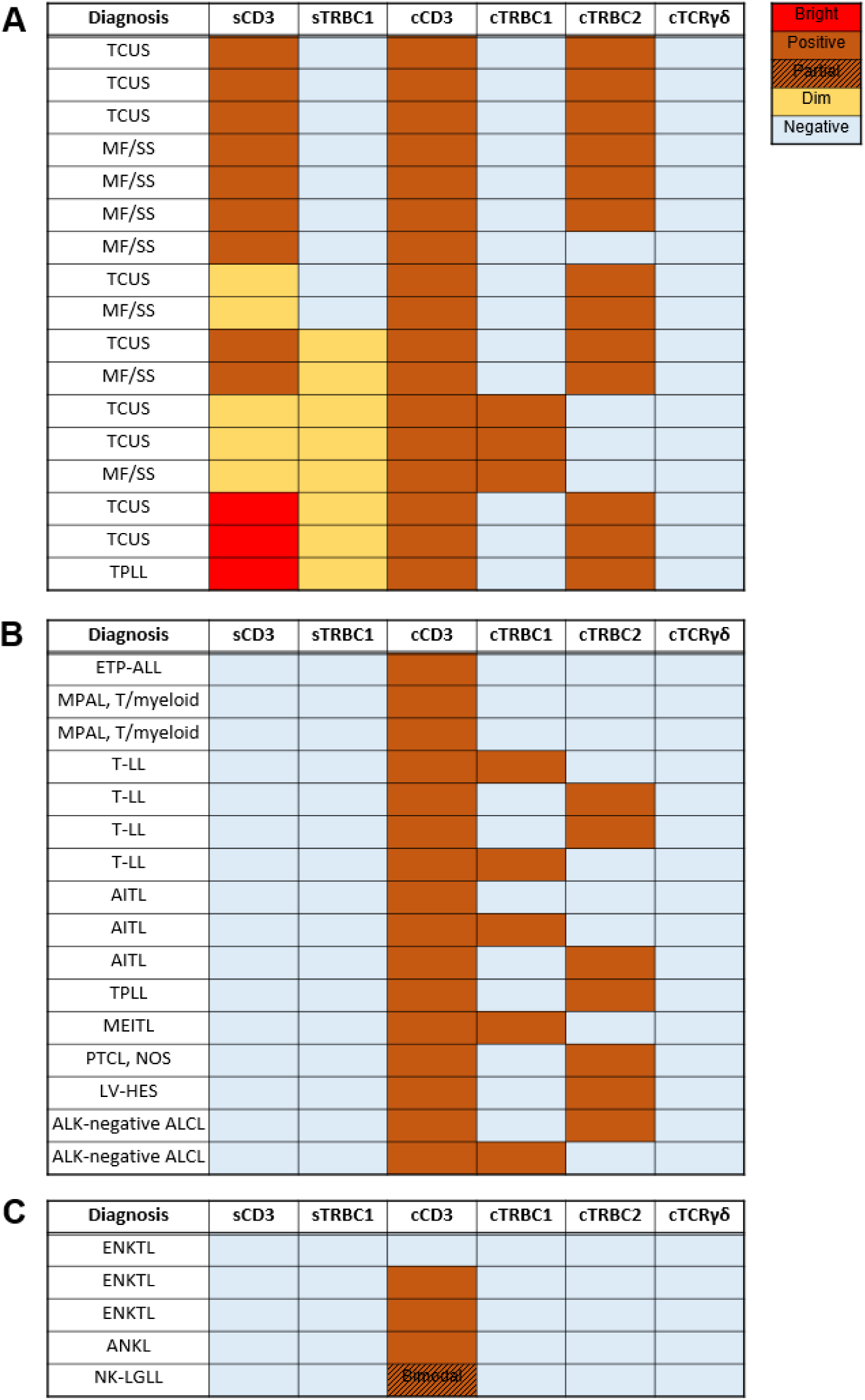
Summary of monotypic T cell populations and NK cell neoplasms profiled by cytoTCR. Expression patterns of sCD3, sTRBC1, cCD3, cTRBC1/2, and cTCRγδ on clonal T cell populations expressing sCD3 (**A**), T cell neoplasms lacking sCD3 expression (**B**), and NK cell-immunophenotype neoplasms (**C**). TCUS, T cell clone of uncertain significance; MF/SS, mycosis fungoides/Sézary syndrome; TPLL, T-prolymphocytic leukemia; ETP-ALL, early T-precursor lymphoblastic leukemia; MPAL, mixed-phenotype acute leukemia; T-LL, T-lymphoblastic leukemia; AITL, angioimmunoblastic T-cell lymphoma; MEITL, monomorphic epitheliotropic intestinal T-cell lymphoma; PTCL, NOS, peripheral T-cell lymphoma, not otherwise specified; LV-HES, lymphocyte-variant hypereosinophilic syndrome; ALCL, anaplastic large cell lymphoma; ENKTL, extranodal NK/T cell lymphoma; ANKL, aggressive NK cell leukemia; NK-LGLL, NK-large granular lymphocytic leukemia.

Notably, one surface CD3^+^ mature T cell lymphoma, which was positive for clonal TCR gene rearrangement by next-generation sequencing (NGS) studies, stained negative for all TCR components including sTRBC1, cTRBC1, cTRBC2, and cTCRγδ (**Figure 3A**), as has been observed previously (Horna et al., 2024a). As expression of surface CD3 by a mature T cell is not possible in the absence of an assembled CD3/TCR complex, we verified surface expression of the CD3/TCR complex using an alternate clone detecting TCRαβ, which demonstrated positive staining (**Supplementary Figure 2**). Positive TCRαβ staining in the absence of TRBC1/2 staining is suggestive of a possible epitope mutation at the site detected by the anti-TRBC antibodies.

We also applied cytoTCR to *n =* 16 T cell populations with absent surface CD3 expression, including 3 T-lineage lymphoblastic leukemias and 13 mature T cell lymphomas. In all mature T cell lymphomas, which each showed clonal TCRβ gene rearrangements by NGS, cytoTCR demonstrated restricted cytoplasmic TRBC expression, simultaneously confirming T cell lineage and clonality (**Figure 3B**).

Finally, we applied cytoTCR to *n =* 5 NK cell neoplasms. Compatible with NK cell lineage, all profiled NK cell neoplasms demonstrated negative staining for all TCR components, but variably showed expression of cytoplasmic CD3 (**Figure 1C-D**; discussed further in **Figure 5**).

### Cytoplasmic TCR expression correlates with developmental stage of T-lymphoblastic leukemia

To explore the developmental significance of cytoplasmic TCR component expression, we applied cytoTCR to a series of seven acute lymphoblastic leukemia specimens with T-lineage immunophenotype. These included two T/myeloid mixed phenotype acute leukemias (MPAL), one early T-precursor acute lymphoblastic leukemia (ETP-ALL), and four T-lymphoblastic leukemias (T-ALL) with a cortical immunophenotype.

All cases of ETP-ALL and T/myeloid MPAL demonstrated dim cCD3 expression without detectable cTRBC1, cTRBC2, or cTCRγδ staining (**Figure 3B**, **Figure 4A**). Interestingly, TCR gene rearrangement studies by NGS on all three cases demonstrated clonal TCRγ rearrangements without evidence of TCRβ rearrangement (data not shown). This pattern corresponds to an early, pre-TCRβ rearrangement stage of thymocyte development, demonstrating concordance between TCR gene rearrangement state and cytoplasmic TCR staining patterns by cytoTCR.

**Figure 4.**
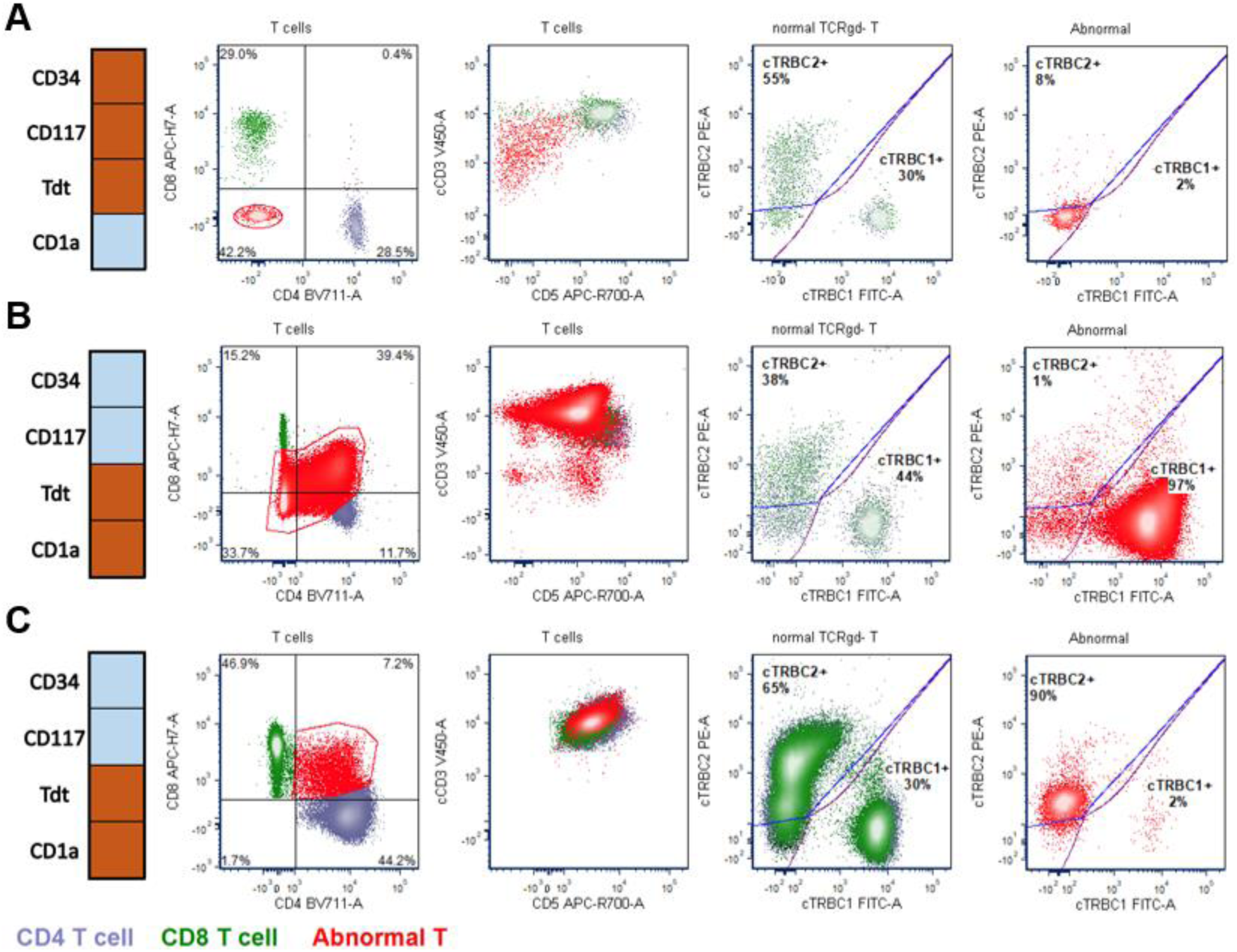
Cytoplasmic TCR expression correlates with developmental stage of T-lymphoblastic leukemia. **A)** A peripheral blood specimen involved by leukemic blasts meeting all WHO5 criteria for early T-precursor lymphoblastic leukemia (ETP-ALL). cytoTCR demonstrates dim cCD3 expression and no expression of cytoplasmic TCR components, compatible with a pre-TCR rearrangement stage. **B-C)** Specimens from two patients involved by T-lymphoblastic leukemia with a cortical immunophenotype, expressing CD1a in addition to partial co-expression of CD4 and CD8. cytoTCR profiling of bone marrow (**B**) and pleural fluid (**C**) demonstrates cCD3 expression in both cases as well as cTRBC1 (**B**) and cTRBC2 (**C**) restriction, respectively, compatible with a post-TCR rearrangement stage. Orange indicates positive staining; blue indicates negative staining.

Conversely, four T-ALL cases, each expressing CD1a and co-expression of CD4/CD8 compatible typical of cortical thymocytes, demonstrated brighter cCD3 expression and overt restriction for either cTRBC1 or cTRBC2 (**Figure 3B**, **Figure 4B-C**). All four cortical immunophenotype T-ALL cases showed clonal TCRβ gene rearrangements by NGS, again demonstrating concordance between molecular studies and cytoTCR.

### NK-immunophenotype NK/T neoplasms frequently express cytoplasmic CD3 in the absence of TCR components

Traditionally, cCD3 positivity has been used as a marker of T cell lineage in the absence of sCD3. However, NK cells are known to cytoplasmically express all CD3 subunits including CD3ε, CD3δ, CD3γ, and CD3ζ, which can form cytoplasmic CD3ε/γ and/or CD3ε/δ heterodimers (Anderson et al., 1989; Lanier et al., 1989, 1992; Phillips et al., 1992; Wilk et al., 2021). While surface expression of CD3 requires the assembly of a CD3/TCR complex and therefore defines T cell lineage, cytoplasmic expression of CD3 does not similarly exclude NK cell lineage. It has previously been proposed that the anti-CD3 antibody clone SK7/Leu-4, by detecting CD3ε/γ and/or CD3ε/δ heterodimers and not free CD3ε chain, could discriminate between T and NK cell lineage neoplasms (Shi et al., 2020). To assess this phenomenon, we analyzed five cases of NK lineage malignancies—three extranodal NK/T cell lymphomas (ENKTL) involving the ethmoid sinus and peripheral blood, one case of aggressive NK cell leukemia (ANKL) involving cerebrospinal fluid, and one case of NK-large granular lymphocytic leukemia (NK-LGLL) in peripheral blood.

In all 4 of 5 cases, cytoTCR identified malignant populations with strong cytoplasmic CD3 expression at levels comparable to those of mature T cells; however, none expressed cytoplasmic TRBC1, TRBC2, or TCRγδ (**Figure 5A–D**). TCR clonality studies by NGS were negative for all cases of ENKTL and the case of NK-LGLL. The absence of functional TCR gene rearrangements by NGS, combined with absent expression of all TCR components, demonstrates that cCD3 expression cannot be reliably used to distinguish T from NK lineage.

**Figure 5.**
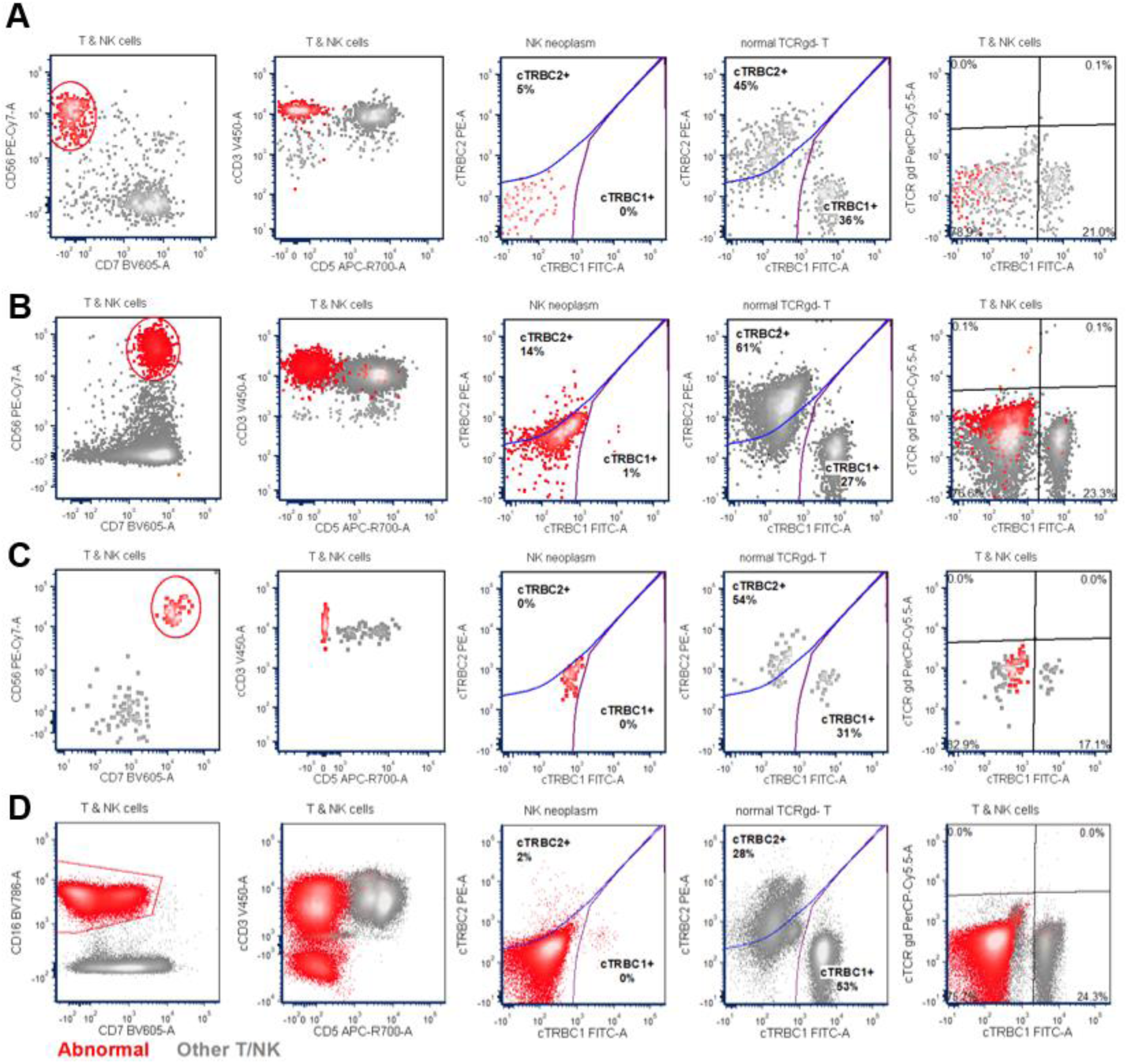
NK/T neoplasms with NK cell-like immunophenotype frequently express cCD3 in the absence of cytoplasmic TCR. **A)** cytoTCR profiling of T and NK cells in ethmoid sinus involved by extranodal NK/T cell lymphoma (ENKTL). **B)** cytoTCR profiling of peripheral blood involved by ENKTL. **C)** cytoTCR profiling of CSF involved by aggressive NK cell leukemia (ANKL). **D)** cytoTCR profiling of peripheral blood involved by NK-LGLL. For all panels: neoplastic populations are circled in red.

### cytoTCR enables efficient confirmation of T cell lineage and clonality in null-phenotype anaplastic large-cell lymphoma

Anaplastic large-cell lymphoma (ALCL) frequently poses a diagnostic challenge because surface CD3 and other conventional T cell markers are often absent, while CD30 is strongly positive. Determining T cell lineage in these cases is essential for classification and to exclude morphologic mimics such as Hodgkin lymphoma, large B-cell lymphoma, or undifferentiated carcinoma with additional immunohistochemical or molecular studies.

We applied cytoTCR to two cases of suspected large cell lymphomas where conventional immunophenotyping was unable to confirm cell lineage. In both cases, standard leukemia/lymphoma immunophenotyping disclosed a CD5^-^CD4^dim^ population with high forward-scatter (**Figure 6A-B**). These populations were negative for myeloid markers (eg. CD13, CD33, CD117, MPO), B-lineage markers (eg. CD19, CD20, CD22, surface and cytoplasmic light chains), as well as surface CD3 expression (data not shown). One case demonstrated CD7 and bright CD2 expression, suspicious for T-lineage (**Figure 6A**), and the other case was predominantly negative for both CD2 and CD7 (**Figure 6B**).

**Figure 6.**
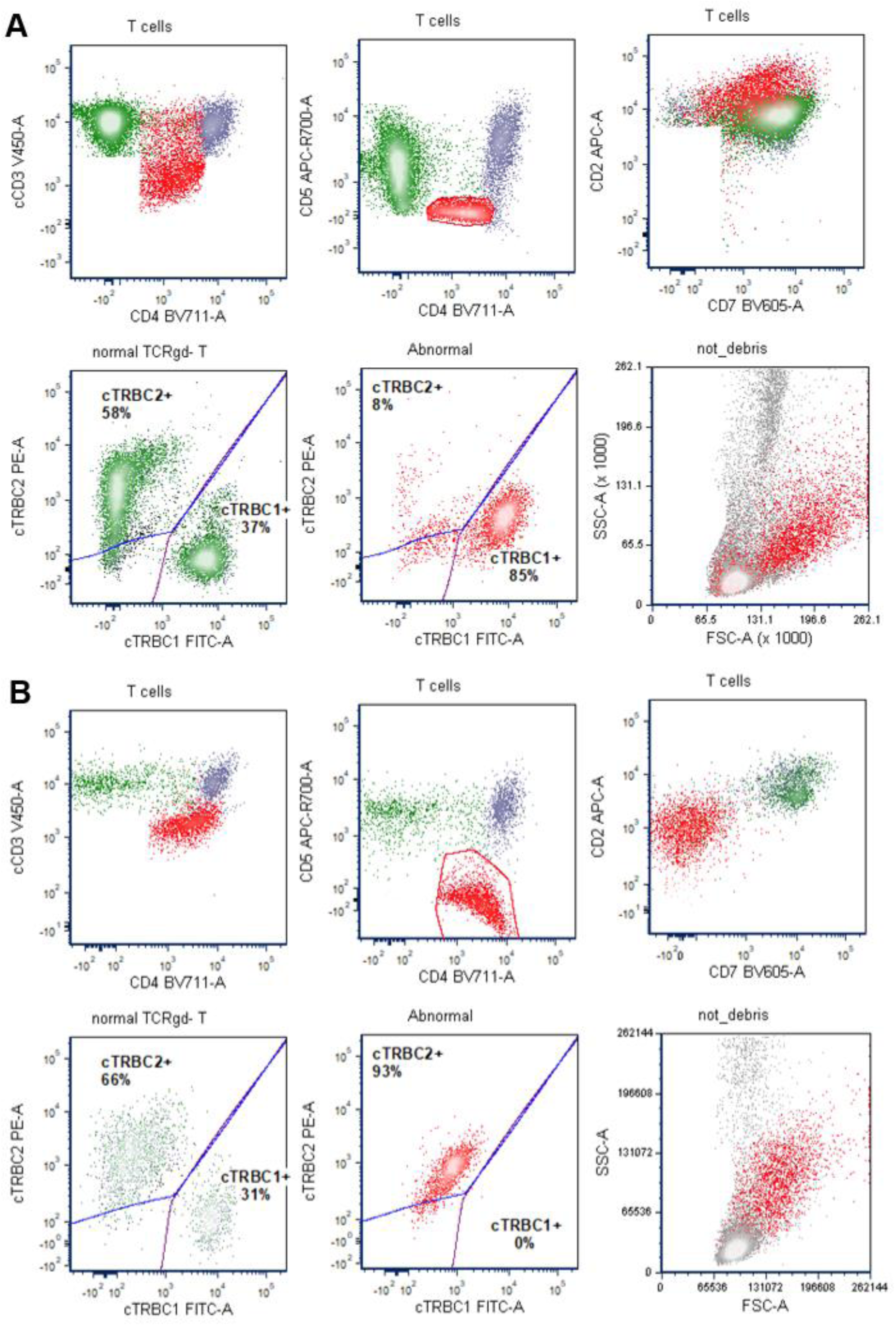
cytoTCR enables efficient establishment of T cell lineage and clonality in anaplastic large cell lymphoma (ALCL). **A)** A submandibular mass with a CD30^+^ neoplasm was profiled by cytoTCR, demonstrating dim cCD3 expression and overt restriction for cTRBC1, compatible with a diagnosis of ALK-negative ALCL. **B)** Bone marrow involved by a CD30^+^ neoplasm of unclear lineage. Based on cytoTCR profiling demonstrating overt restriction for cTRBC2, a diagnosis of null-phenotype ALK-negative ALCL was established. Subsequent T cell clonality studies were positive in both cases.

cytoTCR on both cases demonstrated overt restriction for cTRBC1/2, immediately and simultaneously confirming both T cell lineage and clonality (**Figure 6A-B**). Immunohistochemistry and morphologic review demonstrated an ALK^-^CD30^+^ large cell lymphoma, and TCR gene rearrangement studies were positive on both cases, ultimately confirming a diagnosis of ALK-negative anaplastic large cell lymphoma. These cases illustrate that cytoTCR may expedite the diagnostic assessment of hematolymphoid neoplasms lacking standard lineage-specific cell surface markers.

### cytoTCR enables efficient confirmation of TRBC-polytypic surface CD3-negative CAR-T cells

Chimeric antigen receptor (CAR)-T cells are being increasingly utilized to treat numerous hematolymphoid neoplasms. When used to treat T cell neoplasms such as T-lymphoblastic leukemia (T-ALL), CAR-T products are often engineered to lack canonical T cell antigens such as CD7, to avoid fratricide, and may downregulate surface CD3 expression after activation (Chiesa et al., 2026; Gomes-Silva et al., 2017; Margolskee & Pillai, 2020). These features can make the assessment of residual/recurrent disease in patients who have recently received CAR-T product challenging (Margolskee & Pillai, 2020).

A patient with a history of T-lymphoblastic leukemia who had recently received anti-CD7 CAR-T underwent peripheral blood flow cytometry for disease monitoring purposes. The patient’s leukemic blasts were previously positive for CD7 (bright), CD5, and CD2, and were negative for sCD3, CD4, CD8, CD1a, CD34, and other markers of immaturity. The immunophenotype of the infused CAR-T product was unknown. Peripheral blood flow cytometry demonstrated an abundant atypical T cell population negative for surface CD3 and CD7, with a subset demonstrating increased forward-scatter, but with a normal distribution of CD4 and CD8 (**Figure 7A**). Due to the absent surface CD3 staining, it could not be definitively determined from conventional immunophenotyping alone if this population was clonal. To further characterize this atypical population, we performed cytoTCR on this specimen, which demonstrated the population was polytypic for cTRBC1/2 (**Figure 7B**). cytoTCR therefore enabled efficient exclusion that this population represented residual/recurrent T-lymphoblastic leukemia, and rather likely represented the infused CAR-T product.

**Figure 7.**
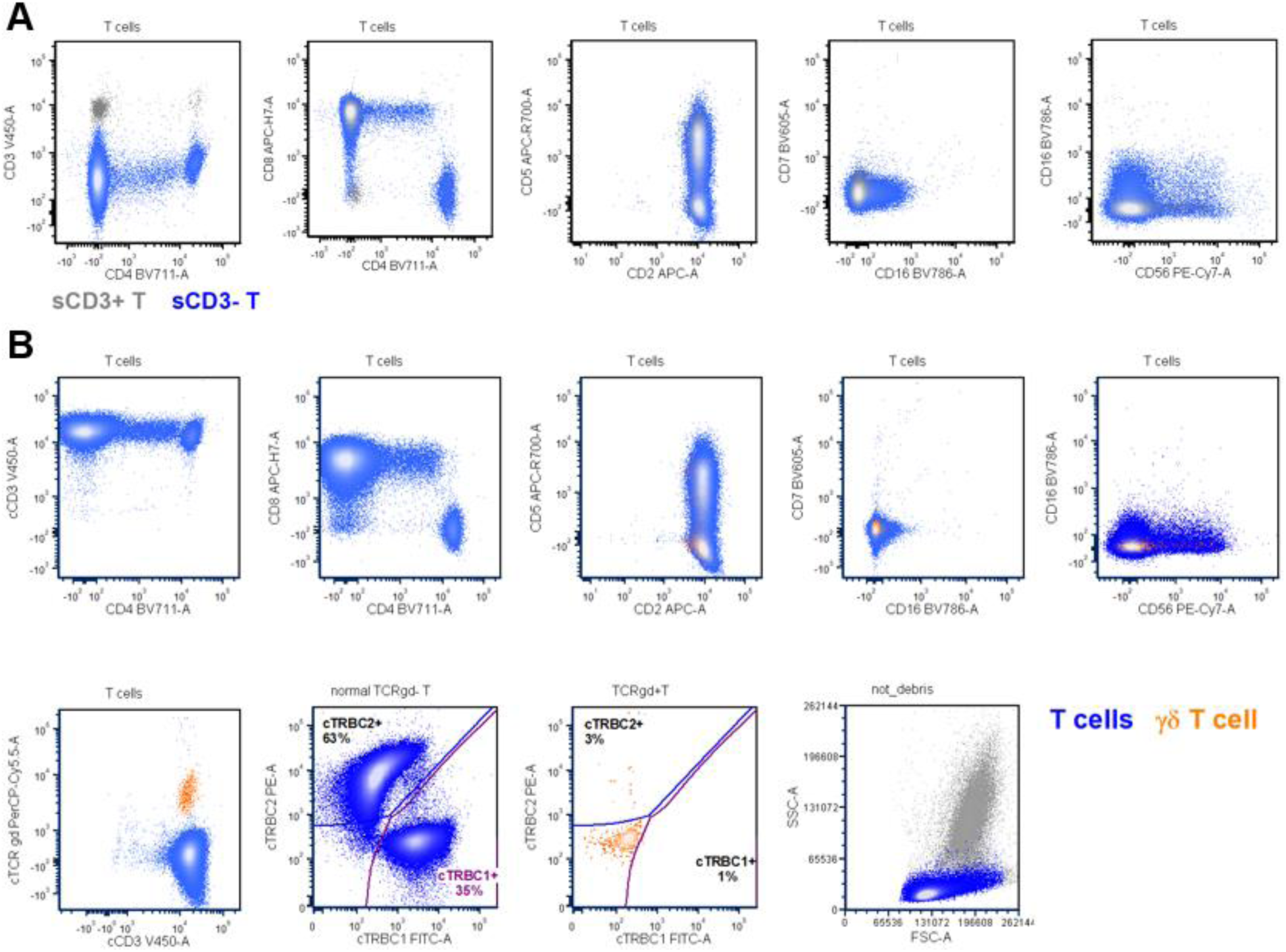
cytoTCR demonstrates that sCD3^-^ candidate CAR-T cell product is TRBC polytypic. A patient with a history of T-lymphoblastic leukemia received anti-CD7 CAR-T cells and underwent peripheral blood flow cytometry immunophenotyping for disease monitoring purposes. **A)** Surface T cell antigen staining reveals a large population of sCD3^-^CD7^-^ events with otherwise normal T cell antigen distributions. **B)** cytoTCR demonstrates the sCD3^-^CD7^-^ population is polytypic for cTRBC1/2, corresponding to candidate CAR-T product engineered with CD7 knockout.

## DISCUSSION

Our study demonstrates that cytoplasmic flow cytometric assessment of all CD3/TCR complex components, including TRBC1/2, TCRγδ, and CD3, offers a powerful tool for both lineage assignment and clonality evaluation in surface CD3-negative T-cell proliferations.

We detect restricted cytoplasmic expression of TRBC1 or TRBC2 in all but one case of T cell neoplasm that were shown to carry TCRβ gene rearrangements by molecular studies. cytoTCR detected restricted TRBC1/2 in these cases of T cell neoplasm independently of surface CD3 expression, and even detected TRBC-monotypic expression in pan-T cell antigen-negative cases of anaplastic large cell lymphoma. Importantly, in T cell neoplasms that demonstrated dim surface staining for TRBC1, we showed that the intensity of sCD3 expression could be used to predict cytoplasmic TRBC1 or TRBC2 restriction, as has been shown previously (Shi et al., 2025). Notably, we identified one case of mature T cell lymphoma that retained surface CD3 expression and stained positive for TCRαβ, but was negative for surface and cytoplasmic staining for TRBC1, TRBC2, and TCRγδ. The absence of detectable surface and cytoplasmic TCR staining in mature T cell lymphomas has been previously described in 4/60 (6.7%) of cases of mature T cell neoplasms (Horna et al., 2024a), and raises the possibility of suboptimal epitope recognition by an anti-TRBC antibody, possibly due to epitope mutation. Therefore, negative staining for cTRBC1/2 and cTCRγδ cannot be definitively used to exclude T-cell lineage, and molecular studies such as TCR gene rearrangement studies by PCR or NGS should be considered for additional evaluation (**Figure 8**).

**Figure 8.**
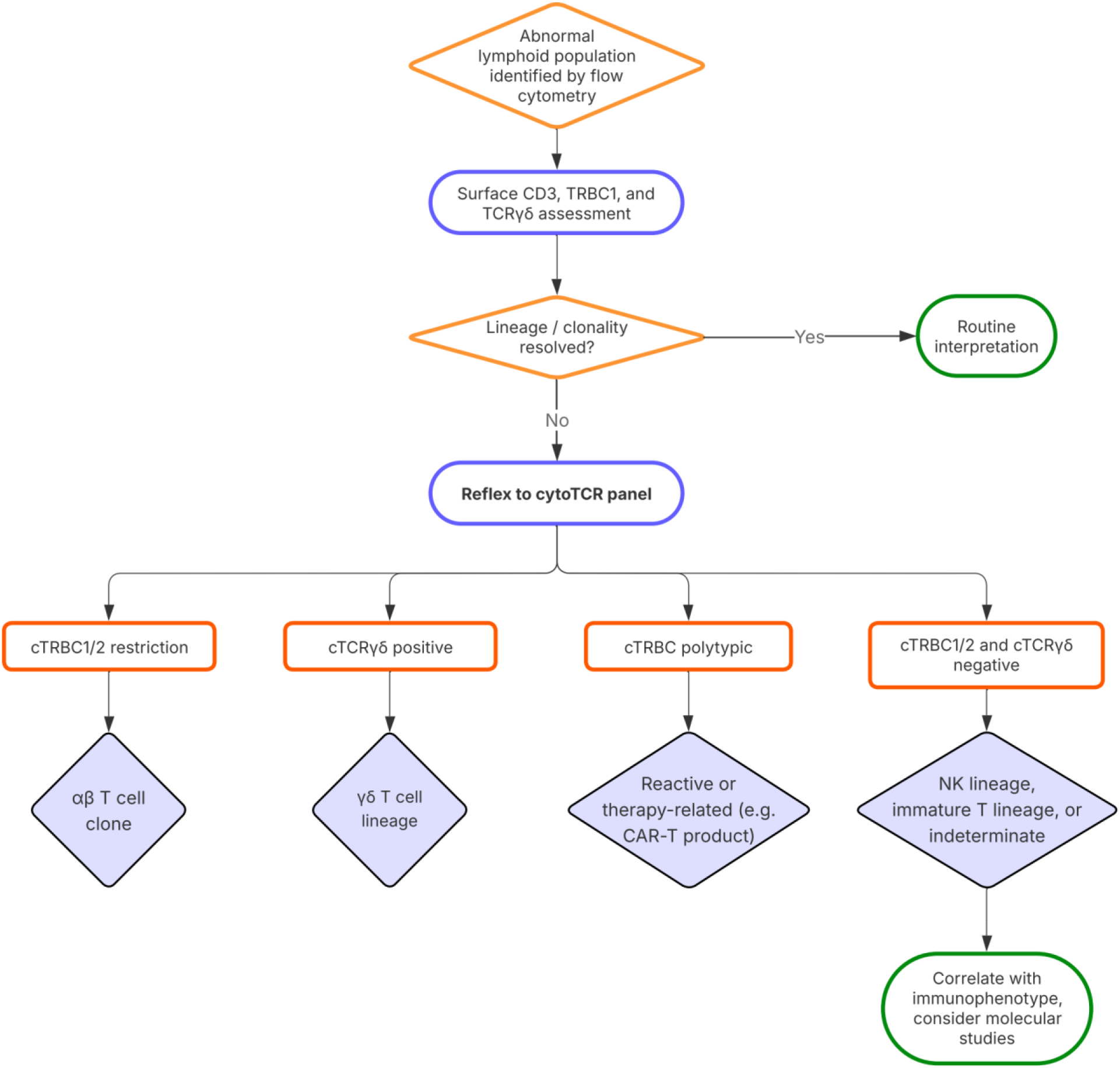
Diagnostic workflow for evaluation of lymphoid populations lacking surface CD3 expression using the cytoTCR panel. Abnormal lymphoid populations are first evaluated using routine surface flow cytometry, including surface CD3 and TRBC1. Cases in which lineage or clonality cannot be established are reflexed to the cytoTCR panel. Cytoplasmic TRBC restriction supports αβ T-cell clonality, cytoplasmic TCRγδ expression supports γδ T-cell lineage, and absence of both supports NK lineage or an indeterminate result requiring correlation with additional studies. Absence of cytoplasmic TRBC expression does not exclude T-cell neoplasia.

One known diagnostic pitfall in flow cytometric evaluation of cytoplasmic CD3 is that natural killer (NK) cells and NK/T neoplasms may exhibit cytoplasmic CD3ε expression despite lacking a rearranged or expressed TCR complex. It has been proposed that the anti-CD3 antibody clone SK7/Leu-4 can be used to discriminate T vs. NK cell lineage by its specific recognition of CD3ε heterodimers (Shi et al., 2020). However, cytoTCR, which also uses SK7/Leu-4 to detect CD3, demonstrated that multiple NK cell neoplasms express cCD3 at levels comparable to background T cells, in the absence of clonal TCR gene rearrangements by NGS. Additionally, in healthy donors without T or NK cell neoplasms, we show there is significant overlap between the cCD3 staining intensity between normal T and NK cells. This is compatible with prior reports that NK cells may cytoplasmically express all CD3 subunits including CD3ε, CD3δ, CD3γ, and CD3ζ, which can form cytoplasmic CD3ε/γ and/or CD3ε/δ heterodimers (Anderson et al., 1989; Lanier et al., 1989, 1992; Phillips et al., 1992; Wilk et al., 2021). Indeed, it is known that reactive NK cell populations, such as NKG2C^+^ “adaptive” NK cells that expand in response to human cytomegalovirus infection/reactivation, can upregulate cytoplasmic CD3 expression (Wu et al., 2021). The difference in results between our study and that of Shi, et al. may be due to differences in other aspects of panel design such as antibody titrations, or different performance of the SK7 clone in the time since the study by Shi, et al. was published (Shi et al., 2020). Therefore, while detection of cytoplasmic TRBC1/2 or TCRγδ may be used to confirm T cell lineage, detection of cytoplasmic CD3 at any intensity is insufficient to exclude NK cell lineage. In routine practice, this distinction is critical because NK/T neoplasms often present with overlapping immunophenotypes and clinical features with T cell lymphomas, yet differ in prognosis and therapeutic approach (Horwitz et al., 2022).

From a practical standpoint, the cytoTCR panel is best used as a reflex assay in cases where routine surface flow cytometry, including surface CD3 and TRBC1, is insufficient to establish lineage or clonality in lymphoid populations that express one or more T/NK antigens or show features suggestive of anaplastic large cell lymphoma (**Figure 8**). This workflow addresses a well-recognized gap in the diagnostic flow cytometry of T-cell neoplasms — namely, the difficulty of assessing T-lineage and clonality when surface CD3 or TCR is absent or down-regulated.

In summary, we demonstrate that cytoplasmic staining of T cell receptor components — specifically TRBC1, TRBC2, and TCRγδ — in conjunction with cytoplasmic CD3 and standard surface immunophenotyping, provides an efficient and robust means to assess T cell lineage and clonality in surface CD3-negative T-cell neoplasms. The cytoTCR panel is applicable across specimen types, aligns with known T cell developmental biology, and enhances the diagnostic power of flow cytometry in challenging cases. Implementation of cytoTCR may reduce reliance on molecular clonality assays and offer more rapid insights in the diagnostic workflow.

## Data Availability

All data produced in the present study are available upon reasonable request to the authors.

## ACKNOWLEDGEMENTS

We thank the Flow Cytometry Laboratory staff at Stanford Health Care for technical assistance and instrument support. Schematic figures were created with BioRender and LucidChart.

## AUTHOR CONTRIBUTIONS

A.J.W. and J.O. conceived of the work and designed experiments. G.G. performed flow cytometry experiments. A.J.W. performed computational analyses. A.J.W. wrote the manuscript with input from all authors.

