## Supplementary Information for "Cytoplasmic staining of T cell receptor components enables efficient assessment of lineage and clonality in surface CD3-negative T cell neoplasms"

**This PDF file includes:**

Supplementary Table 1
Supplementary Figures 1-2

### SUPPLEMENTARY INFORMATION

| Marker | Color | Clone |
|---|---|---|
| CD45 | V500 | 2D1 |
| CD4 | BV711 | SK3 |
| CD8 | APC-H7 | SK1 |
| CD2 | APC | L303.1 |
| CD5 | APC-R700 | UCHT2 |
| CD7 | BV605 | M-T701 |
| CD16 | BV786 | 3G8 |
| CD56 | PE-Cy7 | NCAM16.2 |
| CD57 | FITC | HNK-1 |
| CD3 | V450 | SK7 |
| TRBC1 | PE | JOVI.1 |
| TCRγδ | PerCP-Cy5.5 | IMMU510 |

**Supplementary Table 1.** Flow cytometry antibodies used for surface T cell antigen panel.

**A**

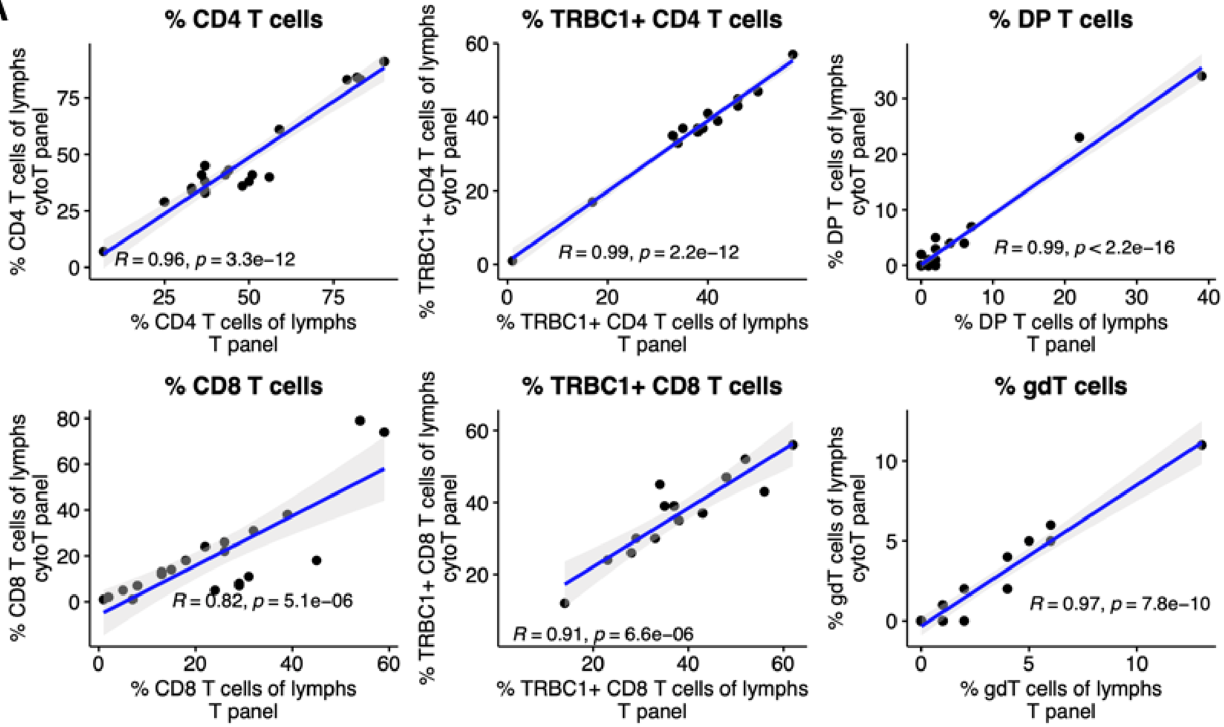


**B**

| Marker | 2.5% | 97.5% |
|---|---|---|
| $cTRBC1^{+}$ | 43.76 | 71.23 |
| $cTRBC2^{+}$ | 27.14 | 52.99 |
| $cTCR\gamma\delta^{+}$ | 0.16 | 12.32 |

**Supplementary Figure 1. Comparison of cytoTCR to surface T cell antigen tube. A)** Peripheral blood specimens from 21 patients without a detectable T cell clone were analyzed by the surface T cell antigen tube (**Supplementary Table 1**) as well as cytoTCR. For all scatter plots, Pearson's *r* and exact two-sided *P* values are shown. **B)** Non-parametric reference intervals for cytoplasmic TCR components, calculated on *n* = 21 peripheral blood specimens without clonal T cell populations.

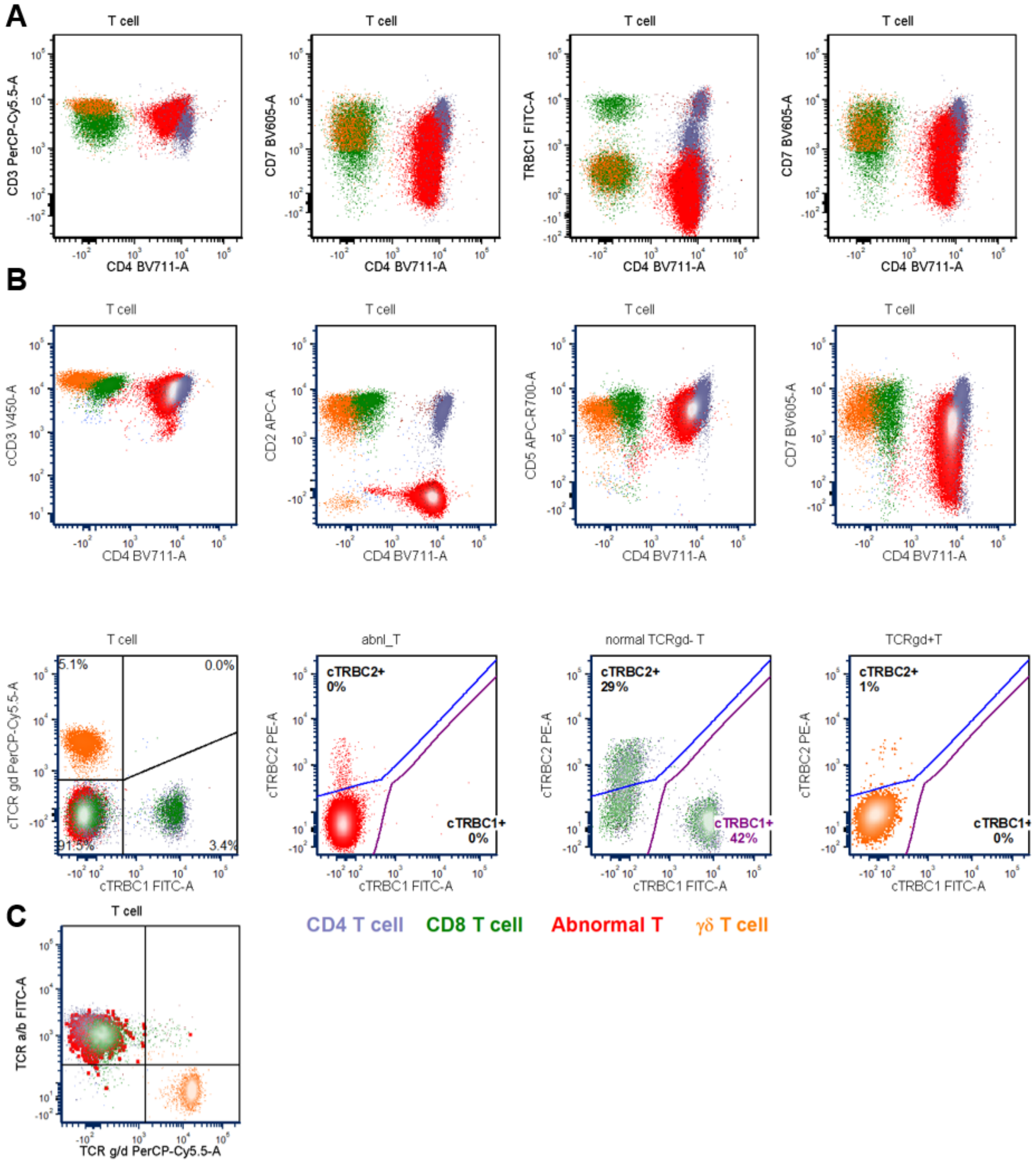


**Supplementary Figure 2. A case of mycosis fungoides/Sézary syndrome with retained surface CD3 expression and no detectable expression of TCR components.** A peripheral blood specimen from a patient with mycosis fungoides/Sézary syndrome was analyzed by the surface T cell antigen tube (**A**) and cytoTCR (**B**). **C)** An additional flow cytometry panel including surface staining of TCRαβ confirms the abnormal population is positive for TCRαβ.
